# What is the Relationship Between Obesity and new Circadian Rhythm Parameters in Turkish Children and Adolescents? A Case-Control Study

**DOI:** 10.1101/2020.09.14.20193938

**Authors:** Mehmet Karadag, Gulay Can Yilmaz

## Abstract

Although the relationship between circadian rhythm parameters and obesity in children and adolescents are widely recognized, there are still not enough studies. The concept of Sleep Corrected Social Jet lag (SJLsc) has been formulated recently and its relationship with childhood obesity has not been established. In this study, we aimed to compare circadian rhythm parameters between obese and normal-weight children and adolescents. Seventy-nine obese and eighty-two normal-weighted children and adolescents aged between 8–17 years participated in this case-control study which took place in Turkey’s Mardin province. Sociodemographic information, Childhood Chronotype Questionnaire, and anthropometric data were collected. The average age of the obese and controls were 12.3 ± 2.3, 12.4±2.2 respectively. Obese young had more evening preference, sleep dept duration, SJL duration, SJLsc duration and morning Evening Scale (MeScale) scores; less mean sleep duration (p<0.005). In regression analyses, BMI z scores were significant correlated with all circadian rhythm parameters, except SJLsc duration, WC z scores were significant correlated with all circadian rhythm parameters, except mean sleep duration. After adjustment, the high mescale scores (OR:1.142, p<0.05) and the presence of a psychiatric disease in the mother (OR:15.075, p<0.05) were associated with obesity. Circadian rhythm parameters can play an important role in the etiology of obesity. Future studies with larger samples and fewer confounding factors are needed to clarify the etiological factors.

## Introduction

Obesity is associated with increased mortality and morbidity. Especially in recent years, obesity has become a serious public health problem all over the world due to its increasing frequency.^1^ The prevalences of overweight and obesity were reported as 15% and 6,6%, respectively, in school-age children in Turkey.^2^ These high rates shows the urgency of etiology and prevention of childhood obesity in Turkey. Excess weight gains, especially in childhood and adolescence, lead to many organic and psychiatric pathologies throughout the rest of life.^3,4^ There are many publications about the etiology of obesity and the evidence base on the different etiologies that contribute to obesity is increasing. In recent publications, circadian rhythm related conditions such as chronotype preferences, social jetlag and sleep debt have also been shown to be a factor in the etiology of obesity.^
5–7
^

Physiological and psychological changes related to circadian rhythm varies between individuals. These differences were defined in the 1900s and were classified as “morning” and “evening” types. Kleitman used the definition “chronotype” for the first time to describe individual differences in circadian rhythm.^8^ Eveningness preference is associated with many diseases such as attention deficit, addiction, cancer and increased metabolic risk.^
9–12
^

Due to incompatibilities between biological rhythm and social-environmental factors, disruptions in circadian rhythm are frequent. This condition is called Social jet lag (SJL).^13^ The effect of SJL on obesity has not a frequently addressed in literature, but it may be an actor in the etiology of obesity.^14^ In a study on adolescents conducted by Malone et al., SJL was found to show a positive correlation with Body Mass Index (BMI) z scores.^6^ It was stated that SJL can be a factor associated with the academic success and cognitive functions of high school students.^15^ In studies conducted with children, it was determined that there was a relationship between sleep habits and SJL and obesity.^16^ However, SJL can be misleading for people who have different mean sleep duration between weekdays and weekends (sleep dept). It may be more appropriate to use the sleep corrected Social jet lag (SJLsc) value for these individuals. SJLsc is obtained from SJL with a calculation to remove sleep duration differences between weekdays and weekends. Thus, the claim that individuals with sleep debt will be calculated better than SJL.^17^

In epidemiological studies, short sleep duration has been shown to be associated with high body mass index and waist circumference. With increased food intake, decreased energy expenditure and changes in appetite-regulating hormone levels such as leptin and ghrelin, sleep deprivation can lead to weight gain and obesity.^18^ Similarly, in a meta-analysis involving 11 longitudinal studies with children, it was determined that long-term shortening in sleep duration doubled the risk of being obese (2.15, 95%CI: 1.64–2.81).^19^

To our knowledge, there is no study in Turkey where all of circadian rhythm parameters are evaluated together in the pediatric age group. In this study, we aimed to compare circadian rhythm parameters such as chronotype preferences, sleep dept duration, SJL, SJLsc and mean sleep duration between obese children and adolescents and normal-weight controls. Our hypothesis was that in obese children, evening preference would be higher, sleep dept duration, SJL, and SJLsc would increase and mean sleep duration would decrease.

## Materials and Methods

### Study Population

The children and adolescents included in the study were selected from among those who applied to the pediatric endocrinology department of the public hospital in Mardin, a city in southeastern Turkey between July 2019 and November 2019. The participants were students studying full-time primary (8.30 am-3.00 pm), secondary or high school education. The obese group was consisted of all obese children who came within this date range and agreed to participate in the study. The control group were consisted of among the children who applied to the same department and did not existence any chronic medical diagnosis and whose body mass index and waist circumference were below 95^th^ percentile. Written and verbal consent was obtained from all the children and adolescents included in the study and their parents. Exclusion criteria have been determined as not attending primary and secondary school regularly in a school in Mardin, the presence of self-reported acute illness, the diagnosis of sleep disorders according to The Diagnostic and Statistical Manual of Mental Disorders-5 (DSM-5) as determined by a child and adolescent psychiatric with a brief psychiatric interview and having a chronic illness that may affect growth and development. Ethical approval was obtained from the Ethics Committee of the University of Health Sciences, Diyarbakir Gazi Yasargil Education and Research Hospital (Date: 14.06.2019 Issue: 285).

Initially, 93 obese and 93 healthy controls were asked to participate in the study. Since 3 obese (2 children diagnosed with Type 2 diabetes mellitus, 1 adolescent use of herbal tea as a sedative for symptomatic control of sleep patterns) and 4 controls (4 children diagnosed a medical disease) did not meet the inclusion criteria; 11 obese and 7 healthy controls were refused to participate to the study, they were excluded from the study. 79 obese and 82 healty control met the inclusion criteria and they agreed to participate in the research. Then, these 161 children and their families were given measuring tools. Finally, the findings include data from 161 children and adolescents. (Figure 1)

**Figure 1:**
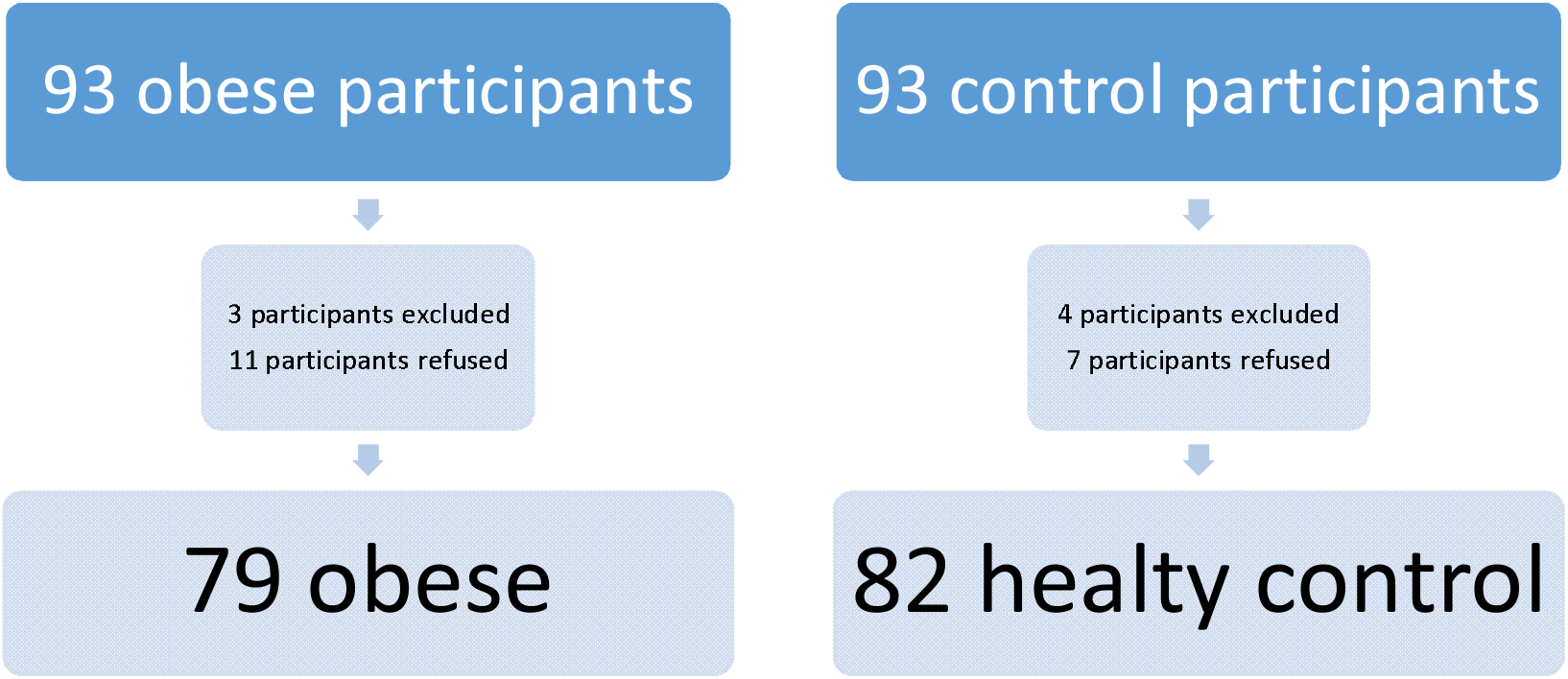
Flow chart for recruitment

### Measurements

*Sociodemographic Data Form*: includes information relating to the child such as gender, date of birth, grade, place of residence, number of siblings, whether she/he applied to a psychiatry clinic. Regarding the family there is information about age, education and occupation. While evaluating the education level of the parents, being a high school and university graduate was rated as higher education level and the rest as low education level. While evaluating place of residence, city center meant Mardin city center; district meant Mardin’s districts; village meant Mardin’s villages.

*Childhood Chronotype Questionnaire (CCQ)*: This questionnaire was adapted from the Munich Chronotype Questionnaire (MCTQ) and the Children’s Morningness Eveningness Scale.^
20,21
^ It is filled out by parents. The first 16 items question sleep/ wake parameters (bedtime, time to turn off lights, sleep latency, wake-up time, time when fully awake, daytime naps). 17–26^th^ items are for determining morningness/ eveningness scores. Items 17, 18, 24 and 25 are scored as reverse. The scores of these 10 questions are summed up. If the total score is 23 or below, it is considered as morningness, if it is between 24–32, it is considered as intermediate form and if it is 33 and above it is considered as eveningness. These parameters were collected under the title Morningness-Eveningness Scale. In addition, to determine the average number of hours the child sleeps, the exact time at which the child sleeps, then exactly what time he wakes up is asked, then the duration of sleep is calculated. If he made a nap during this day, this be added. The same is done for both scheduled days and free days. Then the sleep duration on the scheduled days is multiplied by 5 and the sleep duration on free days is multiplied by 2. Then the sum of the two results found is divided by 7. Thus, the mean sleep duration is obtained. Turkish validity and reliability study for the scale was done by Dursun et al.^22^

SJL and SJLsc parameters were calculated with formulas created in previous studies.^
13, 17
^ SJL is found by the absolute value of the difference between the middle hour of sleep on free days (midsleep on free days- MSF) and the middle hours of sleep in workdays (midsleep of workdays- MSW). The midpoint of sleep is the time between night sleep start time and morning wake-up time. The SJLsc calculation is slightly different. Half of the weekly mean sleep duration are added to both the sleep start time on holidays and the sleep start time on school days, then the values found are subtracted from each other and the absolute difference is found and is named as SJLsc. These can be shown with the formulae; MSFsc = sleep onset on free days + half of the average weekly sleep duration and MSWsc = sleep onset on workdays + half of the average weekly sleep duration. If mathematical simplification is performed, it can be calculated as SJLsc = MSFsc-MSWsc. While calculating the sleep debt, the average weekly sleep duration was subtracted from the average weekday sleep duration and the absolute value of the result was taken.^17^

*Anthropometric measurements*: Anthropometric measurement were carried out in both patients with obesity and controls. Body weight was measured without clothes using “Barimed® Electronic Body Scale SC– 105” which has 0.1 kg precision, and height was measured using the “Ayrton® Stadiometer Model S100” with a sensitivity of 0.1 cm. The Body Mass Index (BMI) was found by dividing the measured weight in kilograms by the square of height in meters.^23^ Using the standards of Turkish children, height standard deviation score (z score), weight z score and BMI z scores were calculated. The waist circumference (WC) was measured at the end of expirium the abdomen was comfortable, the arms were on the side and the feet were united, with a measuring tape that did not stretch when the person was standing.

### Statistical Analysis

IBM SPSS Statistics 25 Package Program was used to analyze the data. Descriptive statistics were given as mean and standard deviation or median and min-max values for continuous data and number and percentage values for qualitative data. In group comparisons, independent groups t-test was used for continuous data, and chi-square test was used for qualitative data. Only in the obese group, two general linear models with the dependent variable being BMI and waist circumference, were obtained. Independent variables were analyzed one by one and variables with p <0.20 (Model 1) were included in the general linear model and a corrected model was obtained. For multivariate analysis, the possible factors identified with univariate analyses were further entered into the logistic regression analysis to determine independent predictors of obese young outcomes. Hosmer-Lemeshow goodness of fit statistics were used to assess model fit. A 5% type-I error level was used to infer statistical significance.

## Results

*Sociodemographic characteristics of the participants:* The mean age of obese children and adolescents was 12.1 ± 2.3 and the mean age of the controls was 12.4 ± 2.2. There was no difference between the two groups in terms of age, gender, education level, place of residence, maternal age, maternal educational status, maternal employment status, number of siblings, father’s employment status, father’s educational status, and father’s psychiatric application history. However, statistically significant differences were found in terms of paternal age, maternal psychiatric application history (**
Table 1
**).

**Table 1:**
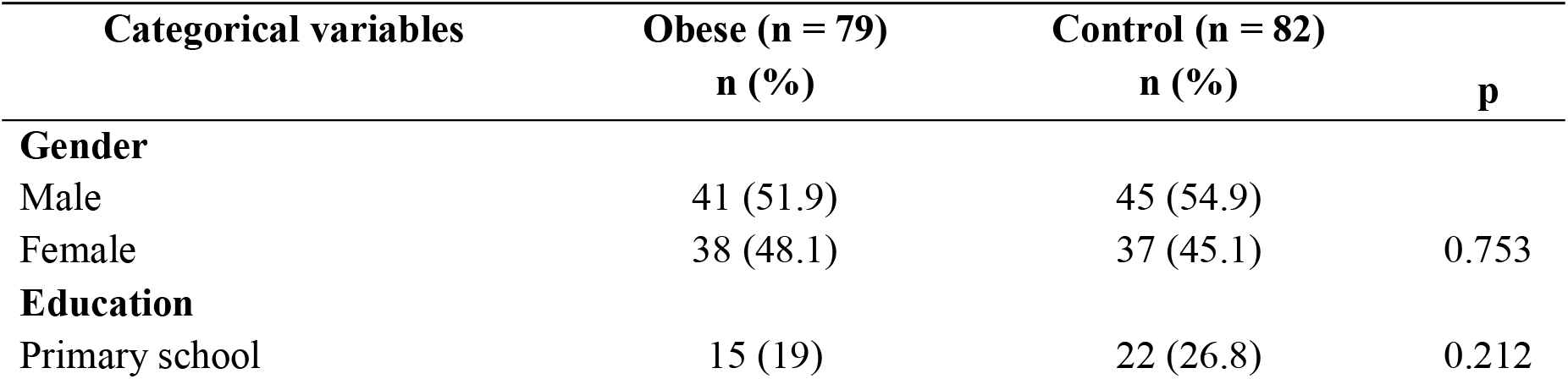
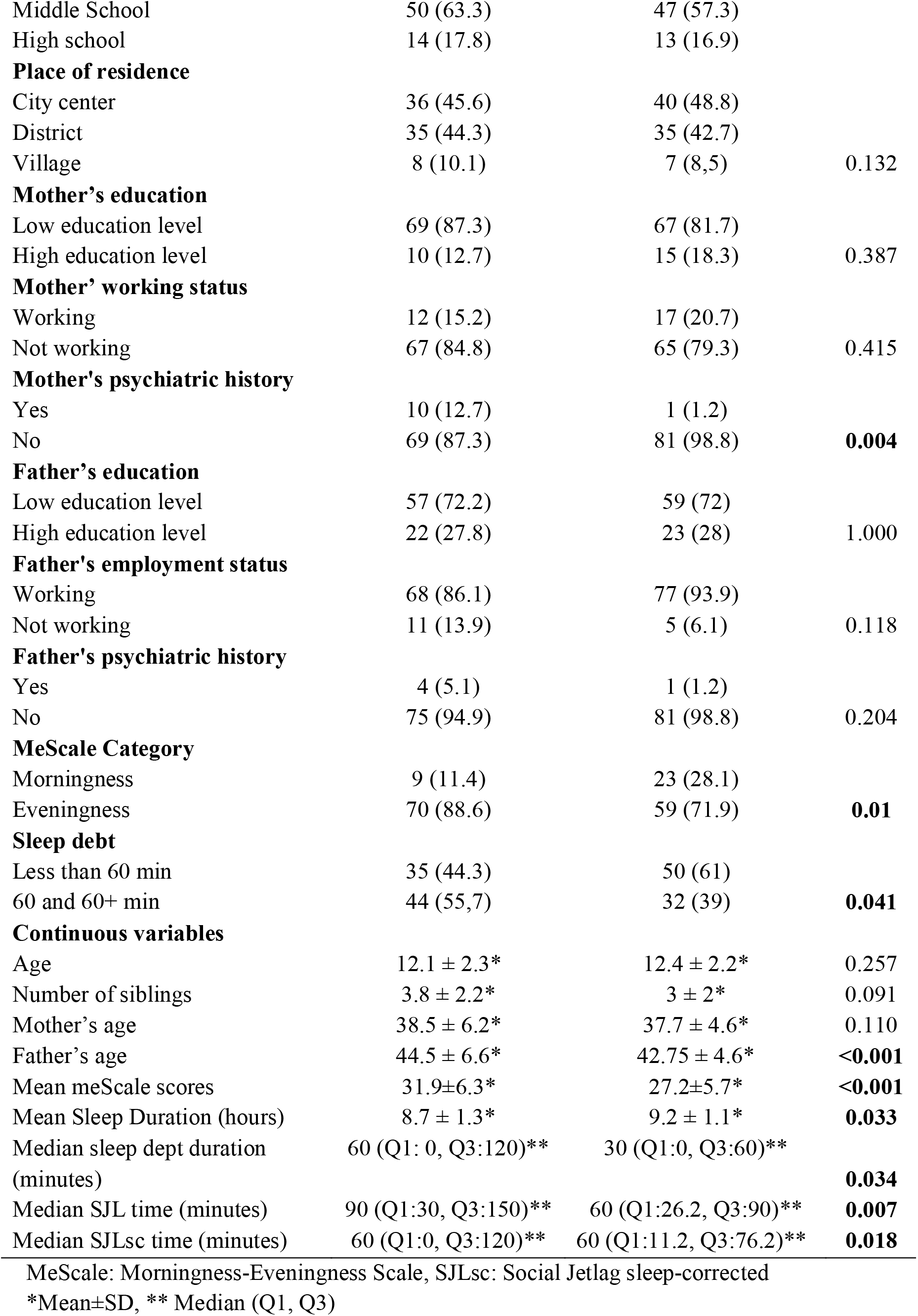
Compare of participants’ descriptive, anthropometrics and circadian rhythm parameters

*Morningness/ Eveningness, Mean Sleep Duration, Sleep Debt and Sleep Corrected Social Jetlag Characteristics of Participants:* In the comparison between obese and health controller, there was a statistically significant difference in terms of all sleep parameters. According to these results, obese young people had more eveningness, less mean sleep duration, higher sleep debts duration, higher SJL and SJLsc durations. **In Table 1
**, comparisons between obese children and healthy controls in terms of sleep parameters are given in detail.

*Anthropometric Characteristics of Participants:* Descriptive statistics about anthropometric characteristics of participants were given in **
Table 2
**. The mean BMI z score of the obese participants was 2.5 ± 0.53 and the mean waist circumference z score was 2.98 ±0.66.

**Table 2:**
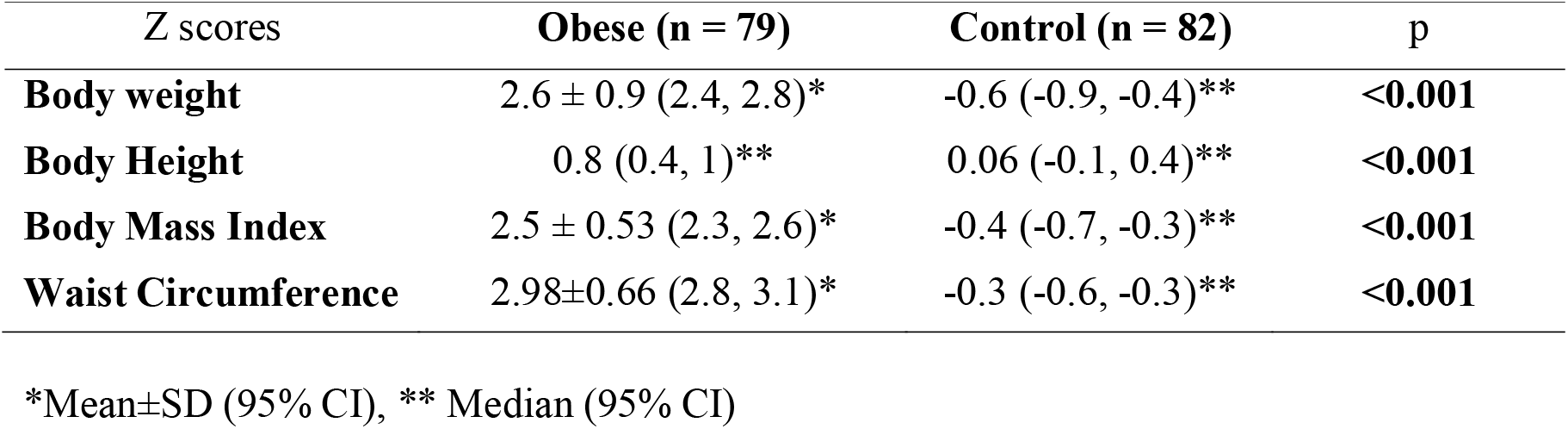
Anthropometric and laboratory data of obese children and adolescents

There were some significant correlations between anthropometric and circadian rhythm parameters. **
Table 3
** demonstrated these difference. BMI z scores were positive correlated with Morning Evening Scale (MeScale) score, sleep dept duration, SJL; negative correlated with sleep duration and no correlated SJLsc. WC z scores were positive correlated with MeScole score, sleep dept duration, SJL, SJLsc, negative correlated with sleep duration.

**Table 3:** Correlations coefficients between circadian rhythm and anthropometric parameters

|  | MeScale scores | Sleep duration | Sleep dept duration | SJL | SJLsc | BMI z scores | WC z scores |
| --- | --- | --- | --- | --- | --- | --- | --- |
| **MeScale scores** | 1 | -0.011 | 0.194* | 0.246** | 0.137 | 0.348** | 0.365** |
| **Sleep duration** | -0.011 | 1 | -0.234** | -0.193* | ,021 | -0.168* | -0.098 |
| **Sleep dept** | 0.194* | -0.234** | 1 | 0.354** | ,062 | 0.246** | 0.220** |
| **SJL** | 0.246** | -0.193* | 0.354** | 1 | ,672** | 0.246** | 0.281** |
| **SJLsc** | 0.137 | 0.021 | 0.062 | 0.672** | 1 | 0.143 | 0.204* |
| **BMI z scores** | 0.348** | -0.168* | 0.246** | 0.246** | ,143 | 1 | 0.880** |
| **WC z scores** | 0.365** | -0.098 | 0.220** | 0.281** | ,204* | 0.880** | 1 |

\*, Correlation is significant at the 0.05 level (2-tailed).
\*\*, Correlation is significant at the 0.01 level (2-tailed).
SJL: Social jet lag, SJLsc: Sleep corrected social jet lag, BMI: Body Mass Index, WC: Waist Circumference

*Regression analyses of related parameters:* **In Table 4
**, Body Mass Index z score of participants were defined as dependent variables, and the predictive factors were investigated including circadian rhythm parameters such as mean sleep duration, morningness/eveningness, sleep debt, SJL, and SJLsc. To create the multivariate model (Model 2), data that were significant at 0.2 alpha level defined in univariate analysis (Model 1) were used. In the univariate analysis, after controlling for confounding variables, it was found that BMI z scores showed positive correlation with number of siblings, mother’s psychiatric history existence, father’s age, and MeScale Scores. The unstandardized beta coefficients values of MeScale Scores were β = 0.082 (0.44, 0.119), p <0.001. However, no significant relationship was found between mean sleep duration, SJL, and SJLsc and sleep debt versus BMI z scores after adjustment.

**Table 4:**
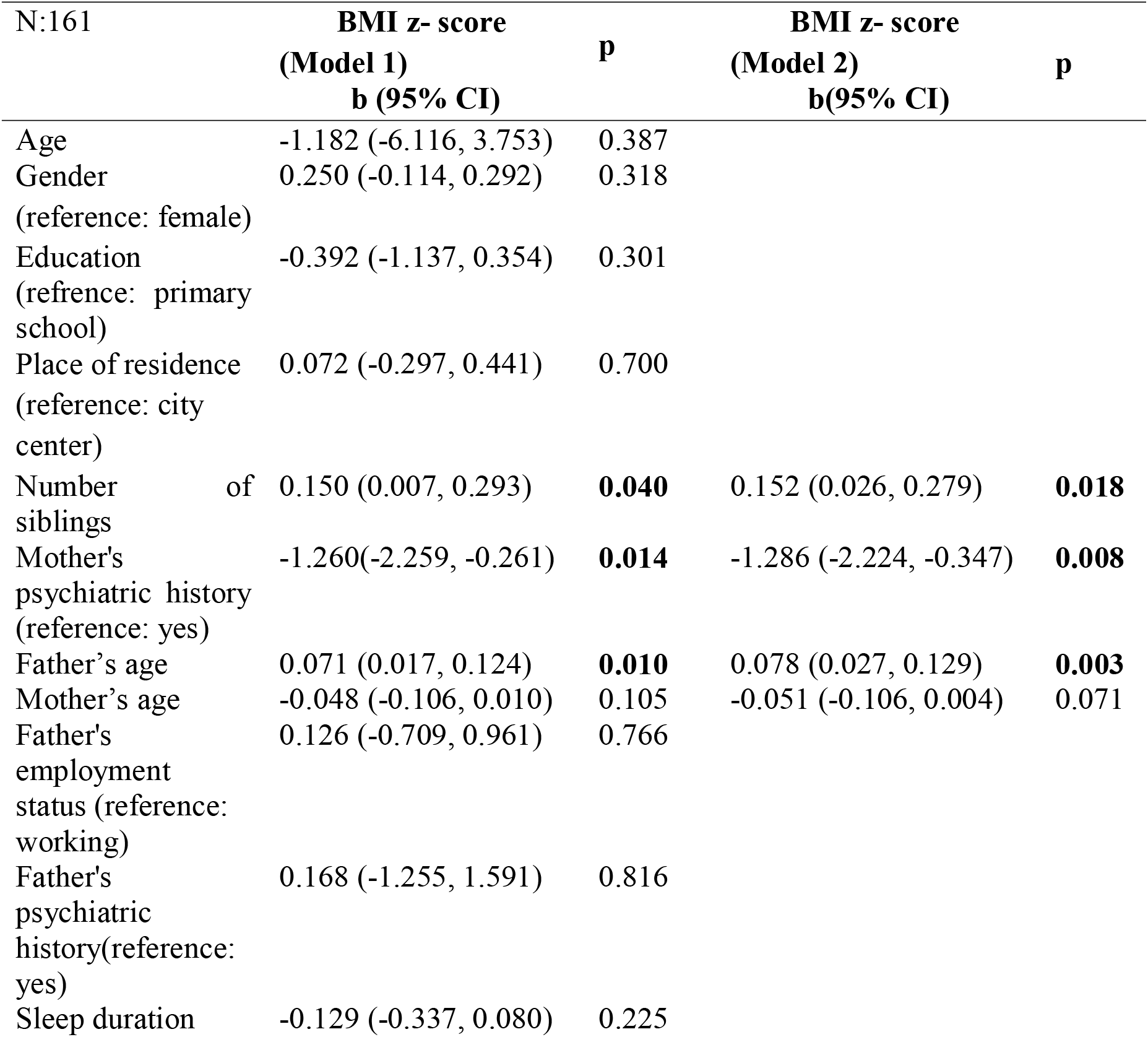
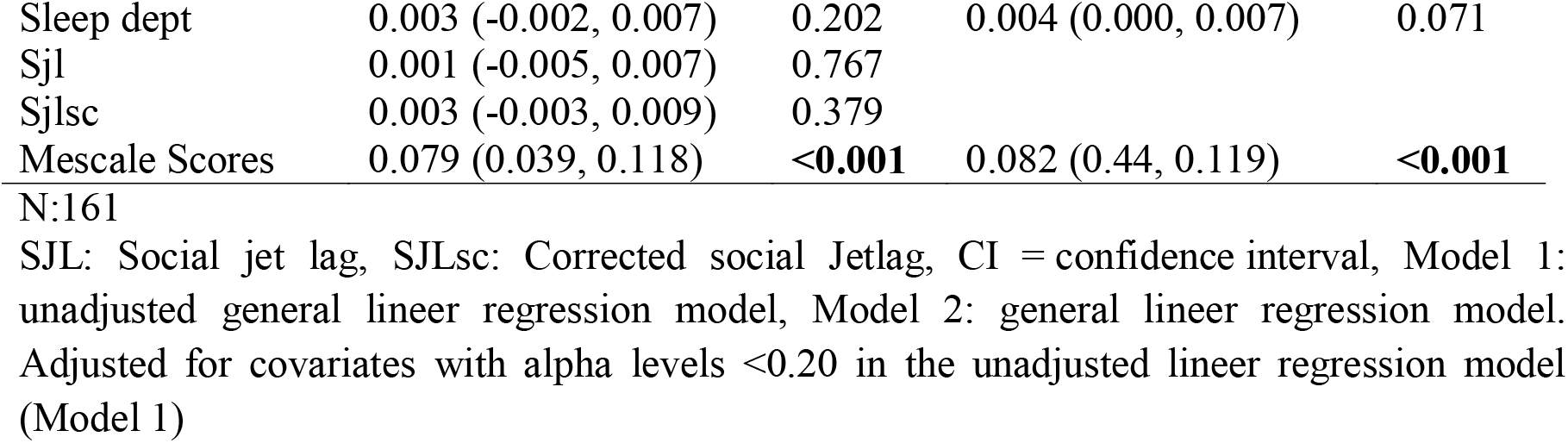
Relationship of chronotype to BMI z-scores.

| N:161 |  | BMI z- score<br>(Model 1)<br>b (95% CI) | p | BMI z- score<br>(Model 2)<br>b(95% CI) | p |
| --- | --- | --- | --- | --- | --- |
| Age |  | -1.182 (-6.116, 3.753) | 0.387 |  |  |
| Gender<br>(reference: female) |  | 0.250 (-0.114, 0.292) | 0.318 |  |  |
| Education<br>(reference: primary school) |  | -0.392 (-1.137, 0.354) | 0.301 |  |  |
| Place of residence<br>(reference: city center) |  | 0.072 (-0.297, 0.441) | 0.700 |  |  |
| Number of siblings |  | 0.150 (0.007, 0.293) | **0.040** | 0.152 (0.026, 0.279) | **0.018** |
| Mother's psychiatric history<br>(reference: yes) |  | -1.260(-2.259, -0.261) | **0.014** | -1.286 (-2.224, -0.347) | **0.008** |
| Father's age |  | 0.071 (0.017, 0.124) | **0.010** | 0.078 (0.027, 0.129) | **0.003** |
| Mother's age |  | -0.048 (-0.106, 0.010) | 0.105 | -0.051 (-0.106, 0.004) | 0.071 |
| Father's employment status<br>(reference: working) |  | 0.126 (-0.709, 0.961) | 0.766 |  |  |
| Father's psychiatric history<br>(reference: yes) |  | 0.168 (-1.255, 1.591) | 0.816 |  |  |
| Sleep duration |  | -0.129 (-0.337, 0.080) | 0.225 |  |  |

|  |  |  |  |  |
| --- | --- | --- | --- | --- |
| Sleep dept | 0.003 (-0.002, 0.007) | 0.202 | 0.004 (0.000, 0.007) | 0.071 |
| Sjl | 0.001 (-0.005, 0.007) | 0.767 |  |  |
| Sjlsc | 0.003 (-0.003, 0.009) | 0.379 |  |  |
| Mescale Scores | 0.079 (0.039, 0.118) | **<0.001** | 0.082 (0.44, 0.119) | **<0.001** |

N:161
SJL: Social jet lag, SJLsc: Corrected social Jetlag, CI = confidence interval, Model 1: unadjusted general linear regression model, Model 2: general linear regression model. Adjusted for covariates with alpha levels <0.20 in the unadjusted linear regression model (Model 1)

**In Table 5
**, the effect of variables with 0.2 alpha significance value in Model 1 on waist circumference z scores of participants was examined by Model 2. After adjustment for the included variables, it was observed that WC z scores showed positive relationship with number of siblings, father’s age, SJLsc duration, and MeScale Scores; negative relationship with mother’s age. The unstandardized beta coefficients values of SJLsc and MeScale Scores were β =0.005 (0.001, 0.01), p=0.025; β=0.101 (0.060, 0.143), p< 0.001, respectively.

**Table 5:**
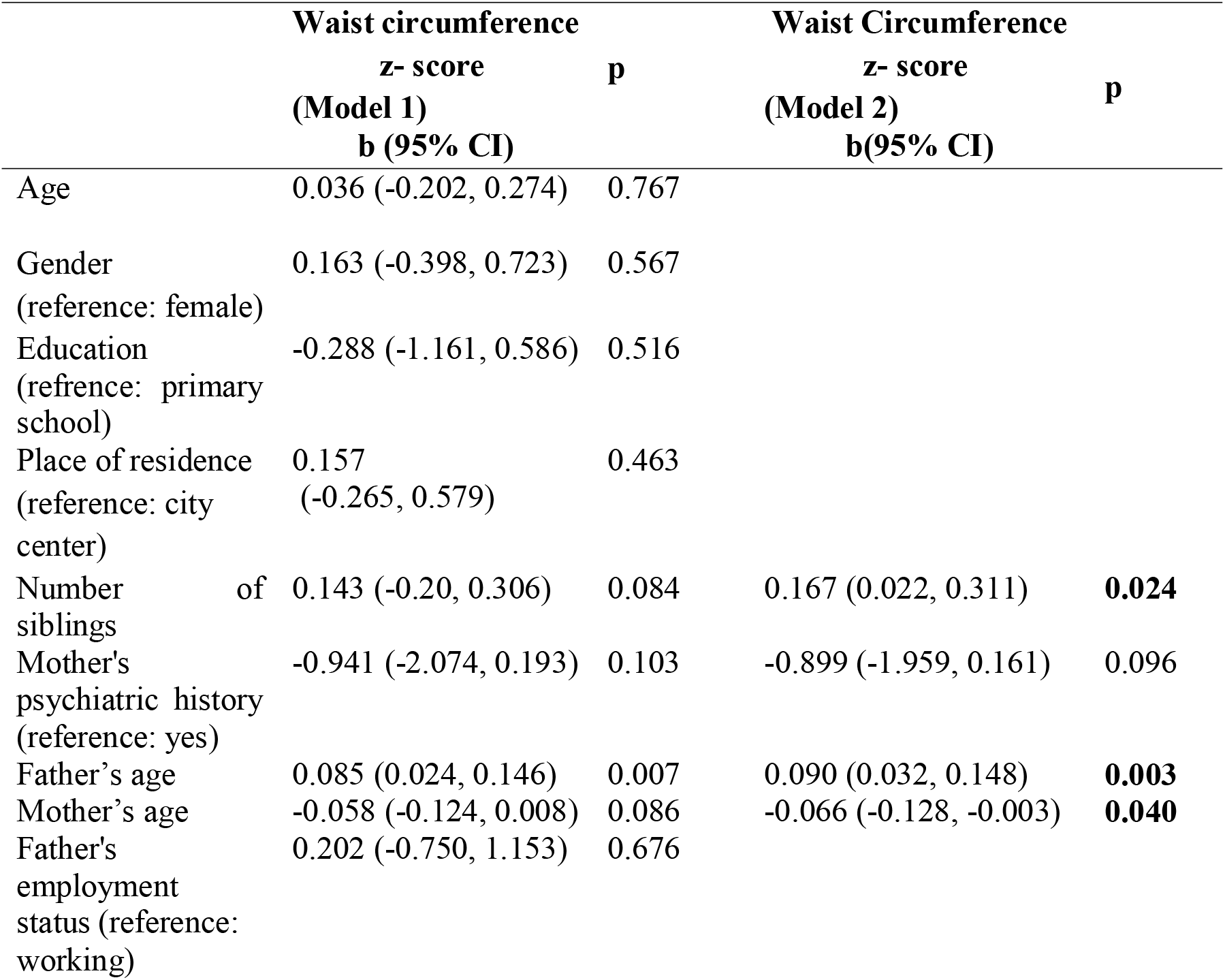
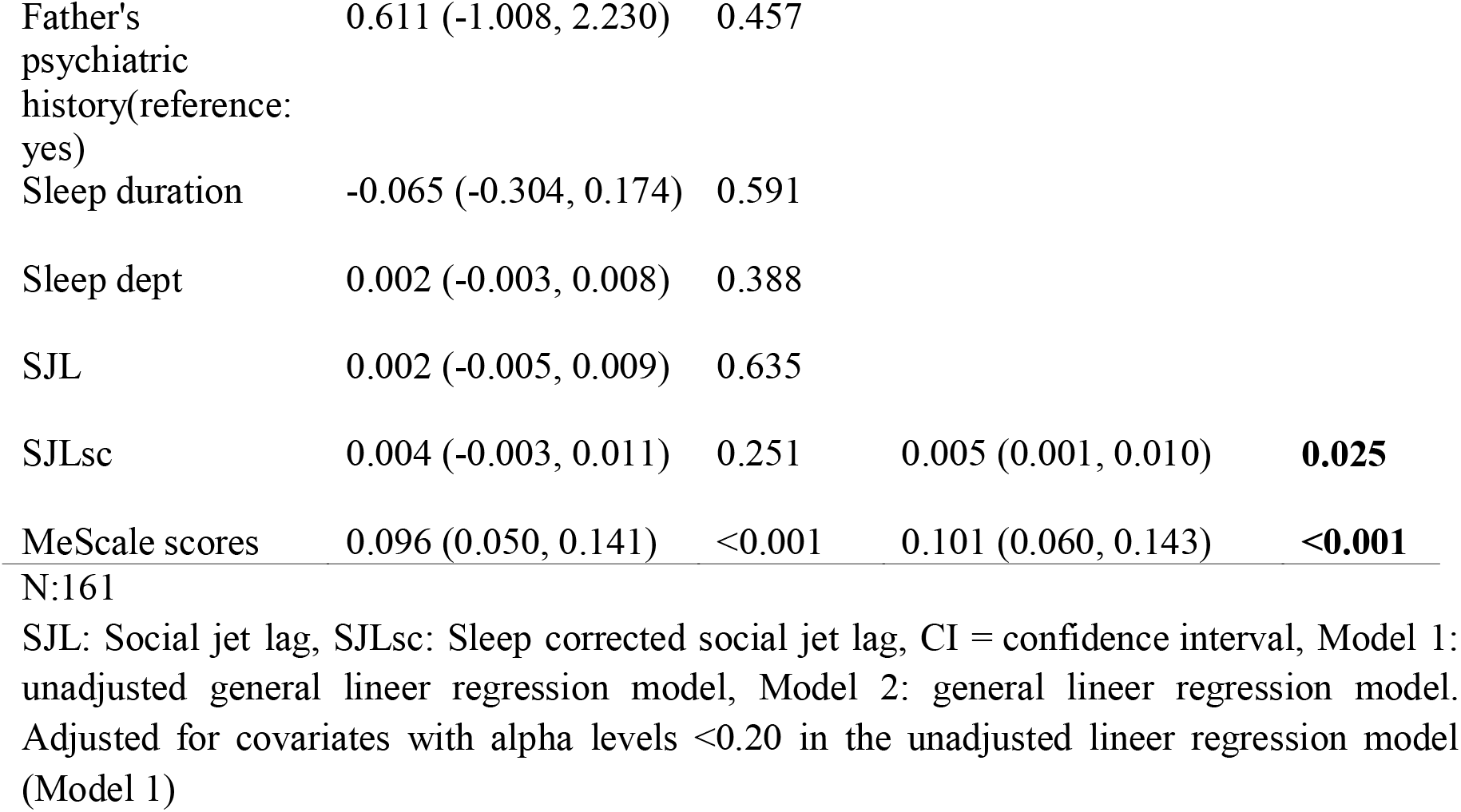
Relationship of chronotype and SJLsc to Waist Circumference z-scores

Logistic regression analysis was carried out by including parameters that may affect obesity, which is the dependent variable (**
Table 6
**). Obesity was positively associated with number of siblings, mother’s psychiatric history existence, and MeScale Scores. These parameters of OR were 1.448, 15.075, and 1.142, respectively.

**Table 6:**
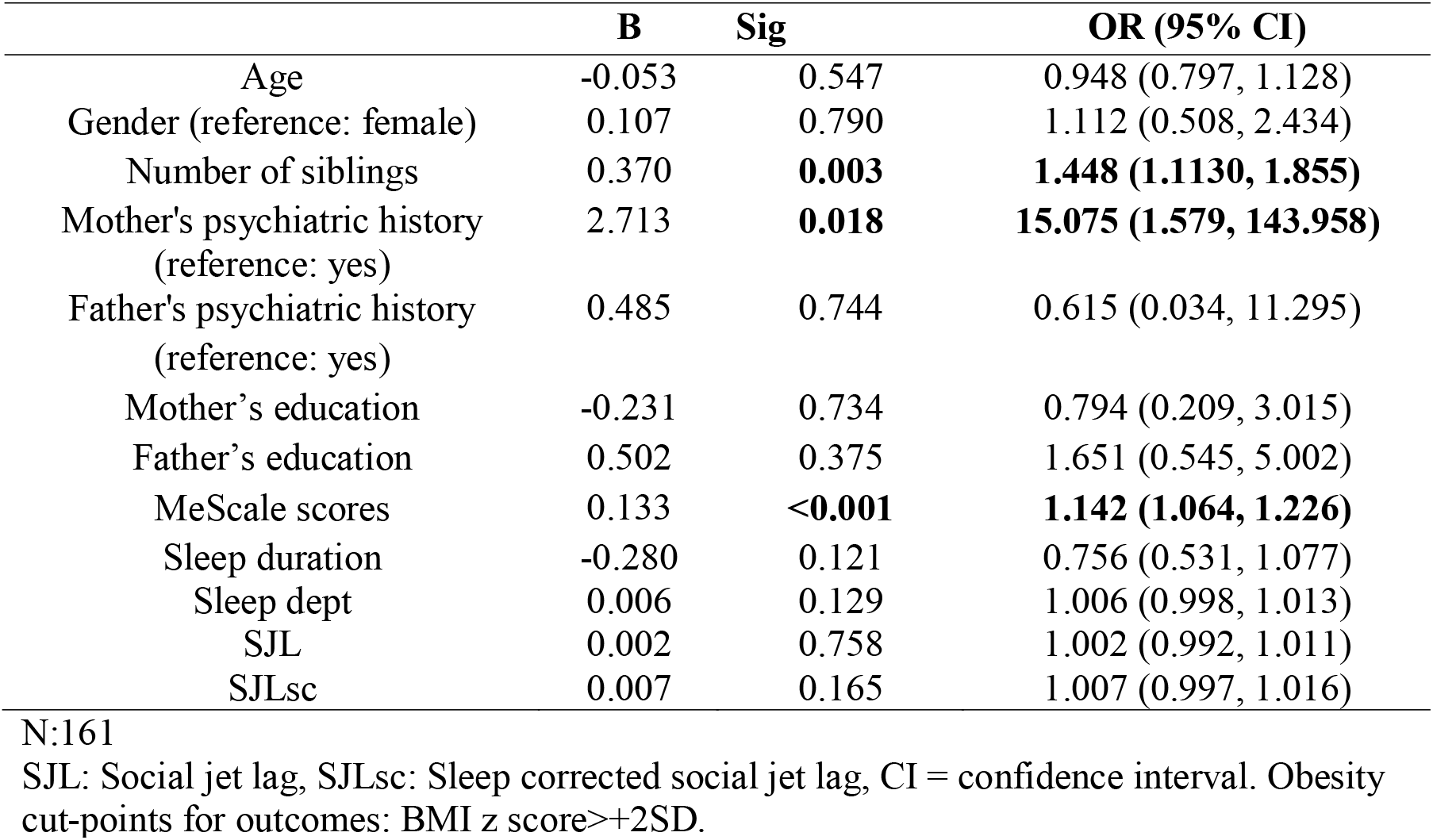
The relationship between obesity and MeScale Scores

|  | **B** | **Sig** | **OR (95% CI)** |
| --- | --- | --- | --- |
| Age | -0.053 | 0.547 | 0.948 (0.797, 1.128) |
| Gender (reference: female) | 0.107 | 0.790 | 1.112 (0.508, 2.434) |
| Number of siblings | 0.370 | **0.003** | **1.448 (1.1130, 1.855)** |
| Mother's psychiatric history (reference: yes) | 2.713 | **0.018** | **15.075 (1.579, 143.958)** |
| Father's psychiatric history (reference: yes) | 0.485 | 0.744 | 0.615 (0.034, 11.295) |
| Mother's education | -0.231 | 0.734 | 0.794 (0.209, 3.015) |
| Father's education | 0.502 | 0.375 | 1.651 (0.545, 5.002) |
| MeScale scores | 0.133 | **<0.001** | **1.142 (1.064, 1.226)** |
| Sleep duration | -0.280 | 0.121 | 0.756 (0.531, 1.077) |
| Sleep dept | 0.006 | 0.129 | 1.006 (0.998, 1.013) |
| SJL | 0.002 | 0.758 | 1.002 (0.992, 1.011) |
| SJLsc | 0.007 | 0.165 | 1.007 (0.997, 1.016) |

N:161
SJL: Social jet lag, SJLsc: Sleep corrected social jet lag, CI = confidence interval. Obesity cut-points for outcomes: BMI z score>+2SD.

## Discussion

The aim of this study was to show relationship with circadian rhythm parameters such as chronotype preferences, SJL, SJLsc, mean sleep duration, sleep dept duration and anthropometric parameters such as BMI and WC z scores in obese children and adolescents and in a normal-weight control group. Our main findings were that obese young had more evening preference, sleep dept duration, SJL duration, SJLsc duration and MeScale scores; less mean sleep duration. In addition BMI z scores were significant correlated with all circadian rhythm parameters, except SJLsc duration, WC z scores were significant correlated with all circadian rhythm parameters, except mean sleep duration. Finally, after adjustment, the high mescale scores and the presence of a psychiatric disease in the mother were associated with obesity. These findings are consistent with a study by Arora and Taheri of 511 adolescents aged 11–13. In this study, evening chronotype and BMI were found to be statistically related. In addition, although this was not evaluated in our study, the frequency of consuming unhealthy snacks, nighttime caffeine consumption, and insufficient daily fruit/vegetable intake were also associated with the evening chronotype in their study.^5^ In studies evaluating social jet, it was observed that obese people had longer social jetlag times in accordance with our study.^24^

In our study, in addition to comparisons between obese and controls, we investigated what factors might affect two important parameters (BMI and WC) that help determine the severity of obesity within the obese group. We found a significant correlation between BMI and WC z scores and circadian rhythm parameters. This finding is in line with our hypothesis. Some results indicated that breakfast skipping, chronotype preference, sleep quality, mean sleep duration, SJL, the midpoint of sleep in free days corrected, conscientiousness, agreeableness and emotional stability are associated factors of dietary patterns. In addition, the risky diet pattern is associated with obesity.^25^

In the analysis of various predictive factors that may affect BMI, it was observed that after correction, eveningness chronotypic preferences and the presence of mother’s psychiatric diseases were associated with increased BMI, but SJL and SJLsc were not. In the regression analysis in which factors affecting the waist circumference were evaluated, circadian parameters such as eveningness chronotypic preferences and SJLsc were statistically associated related after correction. In logistic regression analysis, including confounding factors such as age, gender, the increase in the number of siblings, the presence of psychiatric illness in the mother and high chronotype scores were found to be significant risks in terms of obesity. This is a finding compatible with literature. Most studies have shown that eveningness chronotype and SJL are associated with BMI or WC.^
5,6
^ However, there are studies with different results. In a study conducted in 2018 with pre-diabetic patients, the choice of chronotype and social jetlag were not found be associated with BMI.^26^ Several factors are thought to play a role in this discrepancy. The first and most important of these may be related to the small sample size of studies. If a sufficient number of participants are not included, the results may be unrelated.

Sleep and waking times in adults depend mostly on individual characteristics, therefore evaluations give more accurate results. In children and adolescents, parents play a big role in bedtime and waking hours. Therefore, it is an expected result that there are no statistically significant associations in mean sleep duration and sleep debt between the obese and normal-weight groups.^27^ But there are studies with contrary findings in as well.^
28,29
^ In our study no significant difference was observed between obese and control groups in terms of sleep debt and average sleep duration after adjustment. In addition, some studies have stated that sleep quality, not sleep duration, may be more closely related to obesity.^30^ In our study, sleep quality has not been evaluated, and studies to be designed by considering this difference are needed.

Psychiatric disease in mothers was found to be one of the most powerful markers for obesity. There are different explanations for this situation in the literature. Infancy weight gain during the first year of life, depression and low maternal education were found in a meta-analysis which examined the relationship between environmental risk factors and childhood obesity.^31^ Although it did not reach the level of significance, in accordance with this meta-analysis, the education level of the mothers of the young people in the obese group was lower than controls. Presence of psychiatric disorders in parents is a risk factor for the presence of psychiatric disorders in children.^32^ It is known that the risk of obesity is higher in psychiatric diseases.^33^ This may help to explain the relationship between maternal psychopathology and childhood obesity.

The first limitation is that although the participants are from the same region, information about eating habits and socioeconomic level has not been received. Secondly, it may be a limitation to receive data about sleep parameters via a questionnaire filled out by parents. Using methods such as actigraphy could be a way to increase objectivity.

## Conclusion

As a result, circadian rhythm parameters can play an important role in the etiology of childhood obesity. Especially the eveningness preference and a new circadian parameter named SJLsc, seems to be associated with obesity even after mixing factors are regulated.

## Data Availability

Any data can use in the manuscript.

## Acknowledgements

We would like to thank the authorities who permit and all the participants who attend for this research.

